# Ongoing use of SSRIs and the hospital course of COVID-19 patients: a retrospective outcome analysis

**DOI:** 10.1101/2021.10.25.21265218

**Authors:** Steven H. Rauchman, Sherri G. Mendelson, Courtney Rauchman, Lora J. Kasselman, Aaron Pinkhasov, Allison B. Reiss

## Abstract

**Background:** The SARS-CoV2 virus continues to have devastating consequences worldwide. Though vaccinations have helped to reduce the impact of the virus, new strains still pose a threat to unvaccinated, and to a lesser extent vaccinated, individuals. Therefore, it is imperative to identify treatments that can prevent the development of severe COVID-19. Recently, acute use of SSRI antidepressants in COVID+ patients has been shown to reduce the severity of symptoms compared to placebo. Since SSRIs are a widely used anti-depressant, the aim of this study was to determine if COVID+ patients already on SSRI treatment upon admission to the hospital had reduced mortality compared to COVID+ patients not on chronic SSRI treatment.

**Methods:** A retrospective observational study design was used. Electronic medical records of 9,043 patients with a laboratory-confirmed diagnosis of Covid-19 from 03/2020 to 03/2021from six hospitals were queried for demographic and clinical information. Using R, a logistic regression model was run with mortality as the outcome and SSRI status as the exposure. An adjusted logistic regression model was run to account for age category, gender, and race. All tests were considered significant at p of 0.05 or less.

**Results:** In this sample, no patients admitted on SSRIs had them discontinued. This is consistent with current recommendations. There was no significant difference in the odds of dying between COVID+ patients on chronic SSRIs vs COVID+ patients not taking SSRIs, after controlling for age category, gender, and race. The odds of COVID+ patients on SSRIs dying was 0.98 (95%CI: 0.81, 1.18) compared to COVID+ patients not on SSRIs (p=0.83).

**Conclusion:** In times of pandemics due to novel infectious agents it is difficult, but critical to evaluate safety and efficacy of drugs that might be repurposed for treatment. This large sample size of 9,043 patients suggests that there will be no significant benefit to use of SSRIs to decrease mortality rates for hospitalized patients with Covid-19 who are not currently on SSRI medications. This study shows the utility of large clinical databases in addressing the urgent issue of determining what commonly prescribed drugs might be useful in treating COVID-19.

## Introduction

The COVID-19 pandemic has resulted in an unprecedented worldwide response in the form of a multitude of clinical trials designed to develop efficacious prophylactic and therapeutic interventions (1). The life-threatening global health crisis has encouraged innovation with a focus on rapid results and this has led to the evaluation of existing pharmaceuticals for potential repurposing as COVID-19 treatments (2, 3). The low cost and widespread availability of some drugs already on the market has made them attractive from a social and medical perspective. A number of anti-viral agents are under investigation (4 and 5). Unfortunately, the true efficacy of some of these compounds has not been supported by more rigorous clinical trials (6).

The potential value of selective serotonin reuptake inhibitors (SSRIs), typically prescribed for anxiety, depression and obsessive-compulsive disorder (OCD), has been discussed significantly in the scientific literature (7) and lay press (8).

Numerous in vitro studies have carefully delineated multiple inflammatory pathways in which SSRIs might be beneficial in reducing inflammation (9-12). The key role of inflammation in the progression, morbidity, and mortality of COVID has been well documented in the medical literature and anti-inflammatory effects of SSRIs may underlie their possible protective role in COVID-19 (7, 13).

Early in the pandemic a large French multi-center retrospective study (7) suggested the beneficial role of SSRIs in preventing intubation and death in hospitalized COVID-19 patients. The SSRIs needed to be continued within the first 48 hours of hospital admission. The prior use of these drugs in individuals as outpatients before contracting COVID-19 is not clearly described. There was also a noteworthy exclusion of many patients because of incomplete medical records. A key limitation of this ambitious and important review is the sudden inundation of the French health care system with large numbers of very sick COVID-19 patients. The retrospective nature of the study was intended to encourage more rigorous prospective investigations.

There have since been a number of publications on SSRIs and COVID-19, and these have garnered attention from the lay press. Stories have appeared in the Los Angeles Times and CBS News and the subject was featured on the national television news magazine program “ 60 Minutes”. The importance of exploring the role of SSRIs in COVID was noted in Nature by a co-author of this paper, Steven Rauchman (14). With the presence of effective vaccines, conducting a large prospective clinical trial of therapeutics in the US, Europe, and other nations with large vaccination programs loses feasibility. Simply stated, those fortunate populations cease being a control or treatment group available for potential therapeutic drugs, yet the need for effective therapeutics to prevent suffering and death among a significant part of the world population remains. Vaccines will not reach many less advantaged nations in time and, with new strains, breakthrough cases may emerge in vaccinated populations.

The purpose of this study is to explore the utility of SSRIs in the setting of acute COVID-19 illness, not only as a means to resolve the issue of its effectiveness, but to provide a paradigm for evaluating repurposed drugs and to address the issue of maintenance medication continuation/discontinuation decisions in the acute care setting.

## Methods

This study was approved by the Providence Health and Services IRB as a minimal risk study on March 31, 2021. Providence Health and Services IRB is an electronic IRB serving the 52 hospitals and 1,085 clinics within the large Providence System located in 7 states along the west coast. Providence Health and Services IRB is compliant with U.S. Health and Human Services regulations and requires CITI training and conflict of interest attestations for all investigators. All research studies are required to obtain IRB approval or exemption prior to initiation. There are more than 1700 published studies with Providence Health and Services IRB approval. A retrospective observational study design was used. Therefore, it was determined by the IRB that consent would be waived. Electronic medical records of 9,043 patients with a laboratory-confirmed diagnosis of COVID-19 from 03/2020 to 03/2021 from six hospitals were queried for demographic information, admission date; discharge date and disposition; length of stay; admission diagnoses; medications on admission; co-morbidities; age; gender; ethnicity; admission to ICU; ventilator use; supplemental oxygen; oxygen saturation; discontinuation of antidepressant medications upon ICU admission.

Inclusion criteria: adult patients 18 and over admitted with a diagnosis of Covid-19 and on an antidepressant drug during admission.

Exclusion criteria: patients under 18 years of age, without an admission diagnosis of Covid-19 and not on an antidepressant drug on admission.

Using R 3.6.2 (R Core Team, 2021), a logistic regression model was run with mortality as the outcome and SSRI status as the exposure. An adjusted logistic regression model was run to account for age category, gender, and race. All tests were considered significant at p of 0.05 or less. For inclusion in the statistical model, age category [<18, 18-30, 31-40, 41-50, 51-60, 61-70, 71-80, 81+ years] was recoded into the median year for each age range. One person was dropped from the analysis because their self-reported gender [female, male] was listed as “ unknown”. Self-reported race categories were White or Caucasian, Hispanic or Latino, Black or African-American, Asian, Native Hawaiian or Other Pacific Islander, American Indian or Alaska Native, Other, Unknown. Any person who indicated “ refused to answer” or had missing race information was categorized as “ Unknown” for the analysis.

## Results

Demographic information on our population is shown in Table 1.

**Table 1.**

| Demographic |  | Patients on SSRIs (%) | Patients not on SSRIs (%) |
| --- | --- | --- | --- |
|  |  | N=832 | N=8211 |
|  |  | n (%) | n (%) |
| Gender |  |  |  |
|  | Female | 504 (60.6) | 3646 (44.4) |
|  | Male | 328 (39.4) | 4565 (55.6) |
| Age Group |  |  |  |
|  | >81 | 248 (29.8) | 1623 (19.8) |
|  | 71-80 | 203 (24.4) | 1489 (18.1) |
|  | 61-70 | 181 (21.8) | 1683 (20.5) |
|  | 51-60 | 93 (11.2) | 1458 (17.8) |
|  | 41-50 | 59 (7.1) | 799 (9.7) |
|  | 31-40 | 35 (4.2) | 663 (8.1) |
|  | 18-30 | 13 (1.6) | 455 (5.5) |
|  | <18 | 0 (0.0) | 41 (0.5) |
| Primary Race |  |  |  |
|  | White or Caucasian | 409 (49.2) | 2151 (26.2) |
|  | Hispanic or Latino | 277 (33.3) | 4196 (51.1) |
|  | Black or African American | 43 (5.2) | 493 (6.0) |
|  | Asian | 28 (3.4) | 476 (5.8) |
|  | Native Hawaiian or Other Pacific Islander | 2 (0.2) | 60 (0.7) |
|  | American Indian or Alaska Native | 0 (0.0) | 5 (0.1) |
|  | Other | 61 (7.3) | 683 (8.3) |
|  | Unknown | 12 (1.4) | 147 (1.8) |

The odds of dying do not differ significantly in hospitalized COVID+ patients based on whether or not they are taking SSRIs (Table 2).

**Table 2.** Odds of death in COVID+ patients on continuation of SSRIs during hospitalization

|  |  |  |  |  |
| --- | --- | --- | --- | --- |
| **Variables** | **Crude ORs (95% CI)** | **p-value** | **Adjusted ORs (95% CI)** | **p-value** |
| **SSRI on admission:** |  |  |  |  |
| No | 1 |  | 1 |  |
| Yes | 1.09 (0.91, 1.30) | 0.353 | 0.98 (0.81, 1.18) | 0.828 |
| **Sex:** |  |  |  |  |
| Female | 1 |  | 1 |  |
| Male | 1.34 (1.20, 1.49) | <0.001 | 1.54 (1.37, 1.72) | <0.001 |
| **Age category (years)<sup>a</sup>:** | 1.04 (1.03, 1.04) | <0.001 | 1.04 (1.04, 1.05) | <0.001 |
| **Primary race:** |  |  |  |  |
| White or Caucasian | 1 |  | 1 |  |
| American Indian or Alaska Native | 1.09 (0.06, 7.42) | 0.936 | 2.83 (0.14, 19.91) | 0.359 |
| Asian | 1.27 (1.00, 1.59) | 0.047 | 1.52 (1.19, 1.92) | <0.001 |
| Black or African American | 0.86 (0.67, 1.10) | 0.236 | 1.18 (0.91, 1.52) | 0.215 |
| Hispanic or Latino | 0.86 (0.76, 0.98) | 0.018 | 1.44 (1.25, 1.66) | <0.001 |
| Native Hawaiian or Other Pacific Islander | 0.56 (0.23, 1.15) | 0.148 | 0.92 (0.38, 1.95) | 0.849 |
| Other | 1.12 (0.91, 1.38) | 0.261 | 1.28 (1.03, 1.58) | 0.023 |
| Unknown | 1.52 (1.04, 2.18) | 0.026 | 2.13 (1.44, 3.12) | <0.001 |

CI = confidence interval, OR = odds ratio, SSRI = selective serotonin reuptake inhibitor
a = referent age category is <18 years

There is no significant difference in the odds of dying between COVID+ patients on SSRIs vs COVID+ patients not taking SSRIs. The odds of COVID+ patients on SSRIs dying is 1.09 (95%CI: 0.91, 1.31) compared to COVID+ patients not on SSRIs (p=0.35).

There is no significant difference in the odds of dying between COVID+ patients on SSRIs vs COVID+ patients not taking SSRIs, after controlling for age category, gender, and race. The odds of COVID+ patients on SSRIs dying is 0.98 (95%CI: 0.81, 1.18) compared to COVID+ patients not on SSRIs (p=0.83).

## Discussion

As noted previously, the initial study from France that implicated SSRIs as potential therapeutic tools in COVID-19 was beset by a number of limitations (7). A few other small studies have supported the results from France. In a randomized prospective clinical trial of 152 outpatients with confirmed COVID-19 given either fluvoxamine or placebo, early clinical introduction of fluvoxamine decreased likelihood of clinical deterioration over a 15-day period (15). The authors readily acknowledged the difficulty in recruiting patients. The short duration and small sample size were other limiting variables. The authors recommended larger prospective clinical trials.

A larger clinical trial on fluvoxamine was recently completed in Brazil led by investigators from Brazil and Canada (16). This study appears to support the value of fluvoxamine in early intervention in COVID-19 patients in preventing progression, serious complications and mortality. This study is pending formal publication.

Another important SSRI investigation took place early in the pandemic at a San Francisco Bay Area racetrack (17). A large number of racetrack employees were diagnosed with COVID-19 in a very brief period of time. The racetrack physician offered fluvoxamine to the infected racetrack employees and approximately 50% of the employees took the drug while the remainder declined. There was no treatment available for early intervention in COVID-19 at that point in time besides supportive care. According to this study, fluvoxamine prevented serious clinical deterioration and hospitalization.

The initial inquiry by our group was directed at recreating the original French study. The retrospective review presented here is not a double-blind placebo-controlled randomized clinical trial because that would be costly and difficult to execute in the midst of a pandemic.

A significant segment of the adult population in the United States (estimated at 10-20%) is already taking SSRIs and the rate of antidepressant use has been increasing in the last decade (18). This supports the need to use available data to determine definitively whether patients who are already taking antidepressants fare better or worse than patients not taking such medications. This is especially crucial in more severe COVID-19 cases requiring hospitalization. Our study indicates that, in a population hospitalized for COVID-19, there is no clinical benefit of SSRIs that were being taken before and during admission. Since use of SSRIs for anxiety, OCD or depression seems unrelated to other comorbidities known to affect COVID morbidity and mortality (hypertension, diabetes, heart disease), direct impact of SSRIs on the risk factors for severe COVID-19 is minimal. There is one important caveat and that is the weight gain that may accompany use of some SSRIs (19).

A limitation of our study is that our patient population did not have any patients taking fluvoxamine and this is in direct contrast to the French study. Fluvoxamine has been shown to be a sigma-1 receptor (S1R) agonist with the strongest binding affinity to S1R of all the SSRIs (20). S1R is a chaperone protein located at the endoplasmic reticulum-mitochondrion interface that regulates autophagy, an important process in viral evasion (21). It has cytoprotective and anti-inflammatory properties (22). By activating S1R, fluvoxamine exerts immunomodulatory effects and can reduce cytokine production (23). Other SSRIs do not activate S1R as potently and the absence of a fluvoxamine subset of patients may have affected our results (24, 25).

Another important clinical issue is whether SSRIs are continued or discontinued when a COVID-19 patient is hospitalized or admitted to an ICU. SSRIs are frequently discontinued in the ICU patient (26, 27). This results in adverse ICU outcomes (26, 27). SSRIs are often inadvertently discontinued because they are not considered important in acute disease. This can cause increased agitation among these patients and need for additional sedation, which may in turn depress respiration. There are sometimes relative contraindications to SSRI use in ICU patients. ECG changes and coagulation issues have been noted (28, 29). The ICU physicians in this study did not routinely discontinue the use of SSRIs in patients unless medically indicated. Hospital policy is to continue anti-depressants while patients are hospitalized. The health organization has a Clinical Institutes model that shares expertise and research across the region’s 13 hospitals and affiliated hospitals and this policy was already in existence prior to this study and was not in any manner attributable to this study. As we debate the potential value of such drugs in very sick patients with COVID-19, a collateral effect might be a re-examination of ICU drug protocols for all patients. Among the drug classes with anti-inflammatory properties routinely prescribed in the middle aged and senior populations that merit review are the statins, metformin and antihypertensives, particularly ACE inhibitors (30-32). Perhaps there is synergistic activity in patients on several of these medications. Large retrospective studies of these commonly used drugs would begin to resolve these issues.

## Conclusions

In this retrospective study of 9,043 patients hospitalized for COVID-19 in 6 California hospitals of a large hospital system in the state of California, prior use of SSRIs or SNRIs did not reduce mortality. These drugs were continued during hospitalization and had been started prior to the onset of COVID-19, presumably for a pre-existing psychiatric condition.

This study shows the utility of large clinical databases in addressing the urgent issue of determining what commonly prescribed drugs might be useful in treating COVID-19. The ongoing nature of the pandemic despite the vaccine rollout signals a pressing need to mitigate COVID-19 sequelae and the repurposing of readily available and inexpensive medications has the potential to save lives, particularly because rapid implementation could occur. As a result of this study, the use of the SSRI /SNRI drug class does not hold a particular advantage, but a specific member of this class, such as fluvoxamine may be effective as found in other studies. Further, drug combinations that include SSRIs or SNRIs may exhibit synergy in mitigating COVID-19 severity and the data to make these determinations is likely available within health system electronic records (33).

## Data Availability

All data produced in the present study are available upon reasonable request to the authors

## Acknowledgements

This work was supported by the Quality Department at Providence Southern California Region.

## Declarations

### Ethics approval and consent to participate

This study was conducted under a protocol approved by the Providence IRB. Only anonymized data were used in this study, so consent was not required.

### Consent for publication

Not applicable

### Availability of data and materials

The data are available from the corresponding author upon valid scientific request.

